# A Sew-Free Origami Mask for Improvised Respiratory Protection

**DOI:** 10.1101/2020.09.29.20204115

**Authors:** Jonathan Realmuto, Michael T. Kleinman, Terence Sanger, Michael J. Lawler, James N. Smith

## Abstract

Recently, respiratory aerosols with diameters smaller than 100 *µ*m have been confirmed as important vectors for the spread of SARS-CoV-2. While cloth masks afford some protection for larger ballistic droplets, they are typically inefficient at filtering these aerosols and require specialized fabrication devices to produce. We describe a fabrication technique that makes use of a folding procedure (origami) to transform a filtration material into a mask. These origami masks can be fabricated by non-experts at minimal cost and effort, provide adequate filtration efficiencies, and are easily scaled to different facial sizes. Using a mannequin fit test simulator, we demonstrate that these masks can provide optimal filtration efficiency and ease of breathing with minimal leakage. Because this mask provides greater comfort compared to commercial alternatives, it is likely to promote greater mask wearing tolerance and acceptance.

## Introduction

Airborne respiratory diseases spread through pathogens contained within droplets and aerosols expired during breathing, talking, sneezing, and coughing.^1–8^ Surgical masks and respirators establish barriers between infected and uninfected individuals, thereby reducing the transmission of these pathogens. ^9,10^ During pandemics caused by respiratory pathogens, especially when efficacious pharmaceutical treatments are unavailable, many experts recommend universal masking in public.^11–13^ However, overwhelming demand for respiratory protection can severely strain commercial supplies,^14,15^ requiring some front-line workers and others to use improvised or homemade protection. ^16,17^ A major challenge is to develop an improvised mask that both front-line workers as well as non-experts can produce for ad-hoc respiratory protection. Mask construction usually requires manual labor and sewing machines to produce large numbers in a reasonable amount of time and in general they cannot be reused indefinitely, thus for patient and worker protection they must be replaced frequently. Here, our goal is to design an affordable mask that does not require sewing, combines high filtration efficiency with ease of breathing, minimizes leakage that can dramatically reduced overall mask performance, and provides greater comfort compared to some commercial alternatives thereby promoting mask-wearing tolerance and acceptance.

Improvised cloth masks are widely used by the general public and by essential workers in economically less developed countries; however, little is known about their effectiveness, particularly in a health care setting. ^18^ Surgical masks reduce transmission of droplets from wearer to others (source control)^7^ and can provide some protection from splashed or sprayed body fluids,^10^ but surgical masks do not exhibit adequate filter performance and facial fit characteristics to be considered respiratory protection. ^19^ The efficacy of surgical and cloth masks at filtering severe acute respiratory syndrome coronavirus-2 (SARS-CoV-2), the pathogen responsible for the coronavirus disease 2019 (COVID-19) pandemic, has not been adequately studied, but in a few reports with small numbers of cases, the use of surgical masks reduced the number of infections in healthcare workers. ^20^

Respirators, such as N95 masks, seal around the face to protect the user by filtering contaminated droplets and aerosol particles^10,21^ and are more effective than surgical masks at preventing expelled air leakage during coughing.^22^ Respirators are not typically recommended for the general public or for people not in close contact with infected or suspected patients.^23^ However, given that asymptomatic and presymptomatic transmission of infection has been well documented^24–26^ and some pathogens can remain viable in aerosols for several hours,^27^ mask designs with performance and fit characteristics approaching those of respirators are highly desired.

The effectiveness of improvised respiratory protection is largely determined by the choice of filtration material and quality of design. ^28^ Readily available fabrics, including various cotton fabrics, silk, chiffon, tea towels, and surgical fabric have been repurposed as respiratory filtration systems.^16,17,29–34^ However, even with excellent filtration capabilities, if a mask does not fit properly, virus-containing particles can penetrate through gaps between the mask and face^35^ leading to filtration efficiency decreases of up to 60%. ^30^ The challenge is that good filtration materials are often harder to breathe through, and this higher resistance to flow exacerbates leakage.

In view of the emerging research highlighting the need to protect the public and medical workers from aerosol transmission of COVID-19, the World Health Organization (WHO) recently updated their guidance on the specification and use of face masks.^36^ The WHO’s new guidelines emphasize that, for medical workers, medical grade masks are essential in the treatment of COVID-19 patients, with N95 or equivalent respirators required for all aerosol generating procedures involving these patients. WHO also acknowledges a public need for masks that extends to all settings where physical distancing of at least 1 m is not possible. It goes further to recommend the non-medical mask standard developed by the French Standardization Association (AFNOR Group). The AFNOR specification S76- 001^37^ recognizes that face masks must not only efficiently screen aerosol (minimum 70% filtration efficiency) but also be breathable (maximum inhalation and exhalation resistance of 2.4 and 3 mbar, respectively). AFNOR proposes a 3-layered design that includes an inner absorbant layer next to the face, a middle layer comprised of a non-woven material with optimal filtration properties, and a weather-resistant outer layer. Designs that minimize leakage, such as duck-bill shapes, are also recommended.

Here we present a sew-free, improvised mask design that can be fabricated from a variety of readily available materials including those that can satisfy AFNOR SPEC S76-001. While our design is ideal for general public use, we also envision its use by at-risk, public-facing workers, including emergency services personnel, workers employed in aged care, childcare or education, cleaners, those in the hospitality industry, and public transport and taxi drivers,^38^ and front-line health care works who have a significantly increased risk of infection without adequate access to respiratory protection.^39^ Importantly, our investigation explores filtration properties of various readily available materials and the effect of nominal versus ideal mask fits. Our aim is to provide evidence-based guidance on the selection of materials and the design of improvised masks to enable non-experts the ability to rapidly produce respiratory protection on-demand.

## Materials and Methods

### Origami Fabrication Procedure

The essential components for constructing improvised masks using the origami fabrication procedure are shown in Fig. 1 and include one or more 23 cm square swaths of a chosen filter material, two elastic straps, a nose clip fashioned from a metal wire (e.g., twist tie or coffee bag closure tin), and a stapler and staples. Using a sequence of folds, the filter material is shaped into a mask with staples providing structural support and fastening the elastic straps to the mask. The nose clip helps to seal the mask against the face and is attached to the mask with adhesive or staples. The full fabrication procedure is outlined in Figs. 2 and 3. All mask specimens analyzed in this study were constructed according to the fabrication procedure with the dimensions of each ply measuring roughly 23 x 23 cm and elastic straps measuring 30 cm in length (PADOMA, 1/4 inch wide braided elastic). We used adhesive-backed coffee bag tins for our nose clips (AwePackage Peel-and-Stick Tin Tie - 5-1/2 inch).

**Figure 1:**
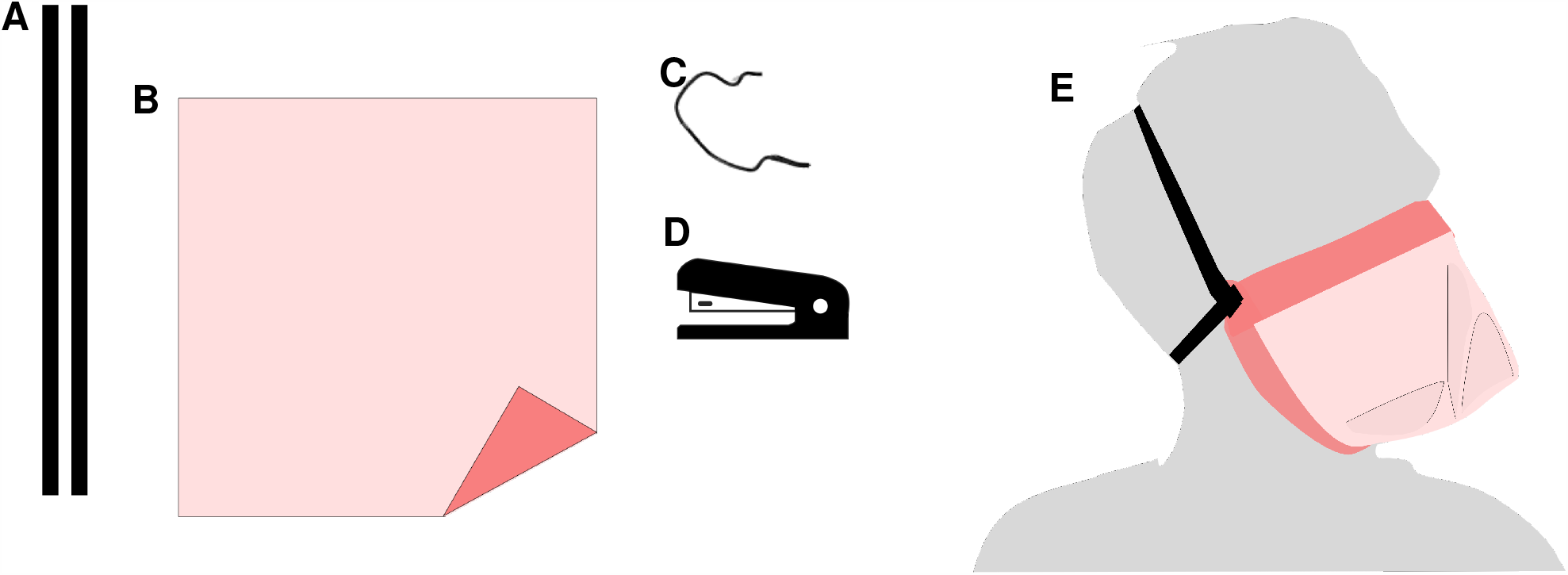
Sew-free origami mask raw materials and tools for fabrication (**A-D**), and illustration of a fabricated mask (**E**). The raw materials needed to fabricate a mask include: (**A**) two elastic straps; (**B**) square piece(s) of filter material(s), which can include multiple plies;(**C**) nose clip material (twist tie, paper clip, or other malleable material); and (**D**) a stapler and staples used to securely join the mask. (**E**) After fabrication the mask is worn over the nose and mouth and secured to the face with the elastic strap.

**Figure 2:**
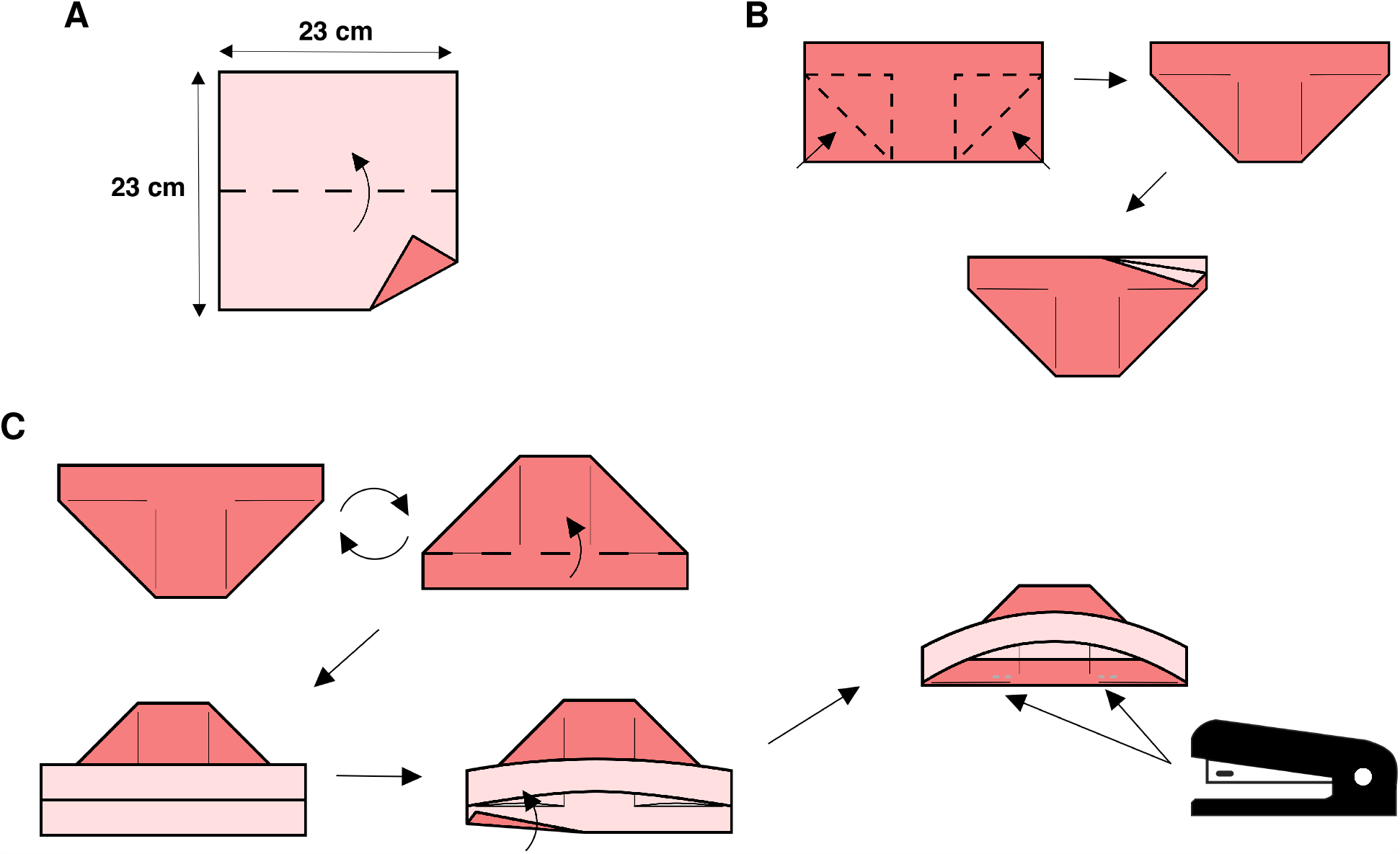
Steps **A**-**C** of the origami fabrication process. (**A**) Fold along horizontal axis. (**B**) Inside-reverse fold each bottom corner leaving an approximate 2.5 cm margin at the top. After the operation you should have a *boat shape* with two *tabs*. (**C**) Rotate piece 180 degrees so you have a *hat shape*. Fold the *top tab* up. Tuck the *bottom tab* into the *hat shape* over the reverse folded corners from Step **B**. Next, staple the folded *bottom tab* to the reverse folded corners.

**Figure 3:**
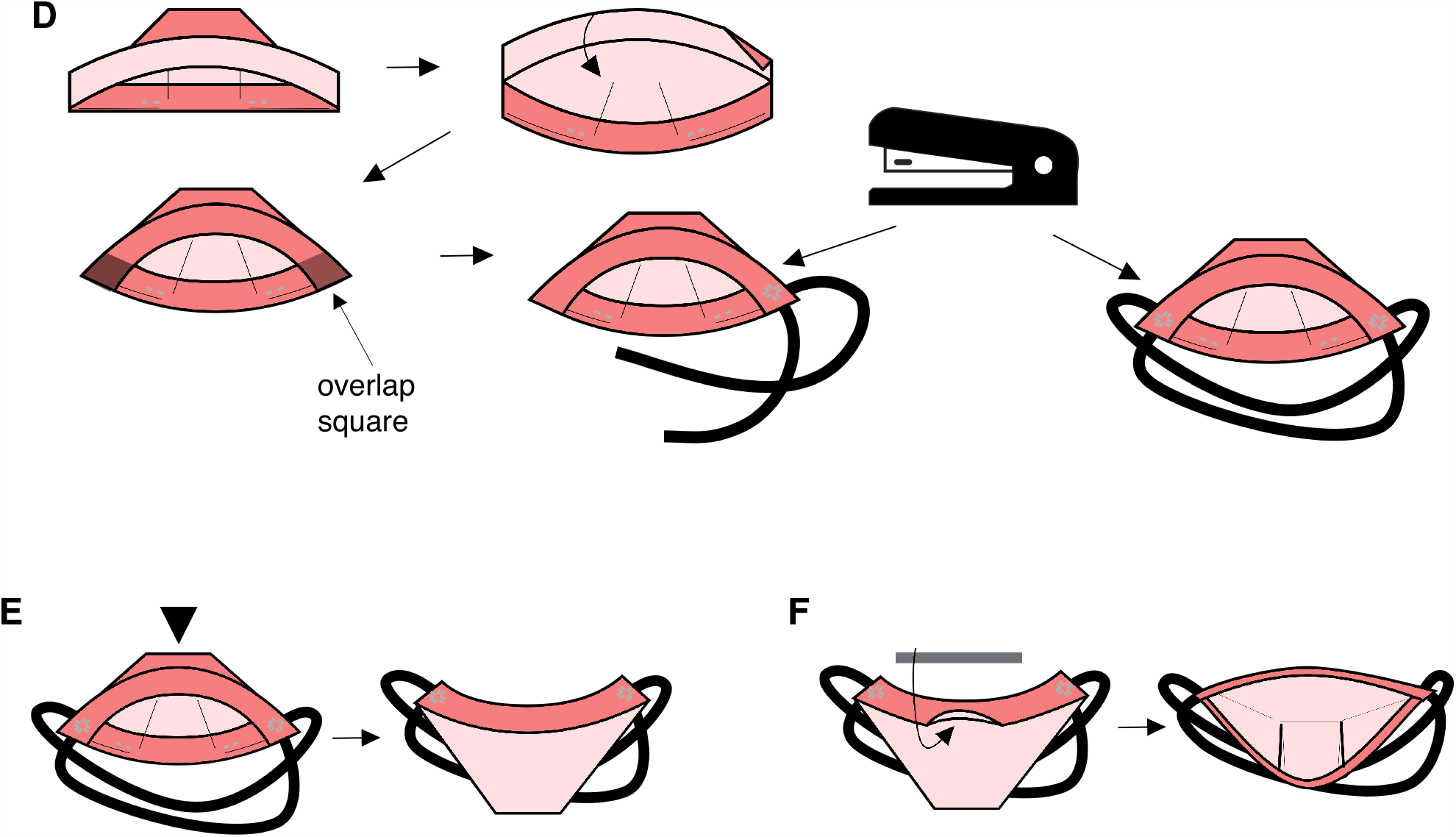
Steps **D**-**F** of the origami fabrication process. (**D**) Fold the *top tab* into the *hat shape* making sure to fold over the *lower tab* so that the edges are aligned and the *tabs* are perpendicular to each other. They should overlap in a square. Starting with one corner, align one elastic strap with one of the folded *tabs* and staple through the mask layers and strap. Repeat for the second elastic strap making sure to align it with the other folded *tab*. Use two more staples to secure the straps. Repeat on the other corner, making sure to align each strap with the correct *tab*. (**E**) Turn the shape inside out. Carefully push the top of the *hat shape* through the folded *tabs*. (**F**) Install the *nose clip*. If using a coffee bag tin, remove the adhesive back, open the *top tab* and place the coffee bag tin sticky side down inside the crease. If using wire, you can secure the wire with a staple.

Inspired by duck-billed style N95 masks, the origami mask is characterized by orthogonal chin and nose folds spanning the perimeter of the filtration area (see Fig. 1 **E**). Commercial masks with similar geometries have been shown to pass standard fit testing using less strap force,^40^ which may provide enhanced comfort. By adapting the dimensions of the chosen filter material, the design can accommodate a range of facial sizes. Some adults have found that masks made from 25 cm square fabric pieces achieve a better fit. Children have found the best fit with square or rectangular material with side dimensions ranging from 15 - 20 cm. For a novice without prior experience, construction takes approximately ten minutes. In our experience, practice decreases assembly time to under five minutes. The origami fabrication technique is a formula for constructing improvised masks that satisfies critical requirements, including ease and speed of assembly, no specialized materials or tools, the ability to accommodate different facial sizes, and the use of readily available materials. The origami fabrication procedure can be used with alternative materials not included here, such as other fabrics with known filtration efficacy (e.g., see ref.^29–34^).

We estimate the total dead space volume between face and mask to be 90 ml based on measurements of accumulated CO_2_ levels inside the mask during normal, resting tidal breathing and assuming an anatomic dead space of 150 ml and a tidal volume of 500 ml. This places the mask below the range of 100 - 165 ml that is estimated for N95 masks. ^41^ Some physiological effects have been noted associated with prolonged use of N95 masks such as increased CO_2_ retention and inhalation from expired breath.^42^ This might be an important consideration for individuals with respiratory or heart impairments. Additional, systematic studies of CO_2_ retention in face masks will be the topic of future research.

### Candidate Filter Media Experiments

The effectiveness of an improvised mask to provide respiratory protection is largely determined by the choice of filtration media. ^28^ To characterize candidate filter media, tests were performed on a variety of materials using the chamber shown in Fig. 4. Aerosol were generated by atomizing an aqueous NaCl solution with a collision-type atomizer (model 3076, TSI, Inc.). The salt concentration within the atomizer was adjusted (nominally 0.2% by mass) to create particles in the 50 – 700 nm diameter range. This test aerosol was introduced into a 1 m^3^ Plexiglas chamber, where it was mixed with dry air to lower the relative humidity (RH) of the chamber air to below the efflorescence point of NaCl (<44% RH). This was done to insure that particles were solid with minimal water content. Perforated inlets and fans were used to insure a homogeneous mixture within the chamber. For tests of the filter media alone, the mannequin head shown in Fig. 4 was replaced with a 47 mm filter holder constructed of perfluoroalkoxy (SKC, Inc.). This filter holder was modified by removing the inlet tube fitting, which allowed the entire front surface of the filter to be exposed to chamber air in order to better simulate mask operation. A second sample line collected chamber aerosol just next to the filter holder for determining the incident particle size distribution. Each of these lines sampled flow at 10 Lpm, corresponding to a 10 cm s^−1^ face velocity for aerosol incident on the filter. Excess flow exited the chamber through a third exit port. A differential pressure gauge (MPXV7002 series, NXP USA, Inc.) measured the pressure drop across the filter material. Selection valves were used to direct either aerosol that penetrates the filter or chamber aerosol into a scanning mobility particle sizer (SMPS).^43^ The SMPS uses a mobility classifier (model 3082, TSI, Inc.) and condensation particle counter (model 1720, Brechtel Manufacturing) to obtain a number-size distribution. A ratio of the size distribution obtained through the sample to that obtained through from the chamber thus provides the penetration efficiency of the filter material over the 50 – 700 nm mobility diameter range.

**Figure 4:**
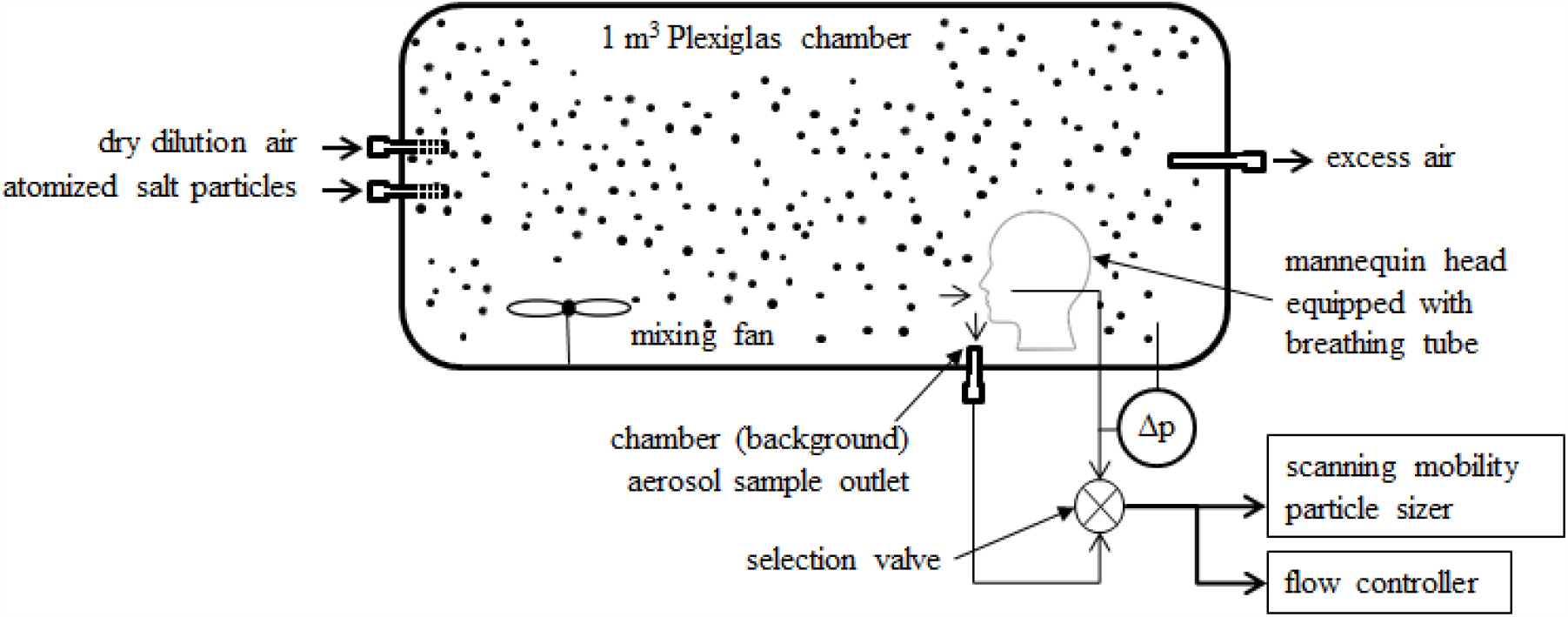
Schematic of the face mask fit test apparatus. A custom-made mannequin head mounted inside a chamber simulates breathing with tubes located at the nostrils and connected to a flow controller. Dried, aerosolized NaCl particles are injected into the chamber and mixed. To test a mask, a specimen is attached to the mannequin face in either the sealed or unsealed test condition. The particle size distributions are monitored inside the chamber and from the inhalation stream of the mannequin with a scanning mobility particle sizer. A differential pressure sensor monitors the pressure drop across the mask.

### Mask Material Selection

Origami masks were constructed using selected filter materials for further analysis, including multiayered “hybrid” configurations. The list of materials used for the origami masks are provided in Table 1, with a more detailed description of each material provided in Table S1. We focused on materials that could be easily and rapidly sourced locally, e.g., from a hardware or craft store. The one exception is the Filti material, which is specifically advertised as a filtration medium for masks and can be sourced only through the manufacturer. Hybrid masks satisfy AFNOR SPEC S76-001 construction standards with the following notable exceptions. (i) The materials we tested employ a hydrophobic inner layer (non-woven polypropylene, NWPP) instead of a recommended hydrophilic one. While it is possible to use three unique materials to construct the mask by selecting a cotton inner layer, we do not expect the filtration performance to change as a result of this small modification. (ii) The materials are not machine-washable according to the AFNOR standard. To the latter point, since these masks are easily constructed using inexpensive materials, we recommend these as disposable (single-use) masks.

**Table 1:** Name and label of each test masks. Additional information on the fabrics and filter media are provided in Table S1.

| Name | Label |
| --- | --- |
| 1-ply Filti | 1F |
| 2-ply Filti | 2F |
| 1-ply Wypall X80 | 1X |
| 2-ply Wypall X80 | 2X |
| 1-ply Muslin | 1M |
| 2-ply Muslin | 2M |
| 1-ply NWPP | 1P |
| 2-ply NWPP | 2P |
| oil towel | P |
| outer ply Halyard H600 | oH |
| inner ply Halyard H600 | iH |
| both plies Halyard H600 | 2H |
| Hybrid 1 (1P - 1F - 1P) | H1 |
| Hybrid 2 (1P - MERV13 - 1P) | H2 |
| Hybrid 3 (1P - MERV8 - 1P) | H3 |
| KN95 Mask | K |
| Surgical Mask | S |

## Fit Test Simulator Experiments

The experimental apparatus in Fig. 4 was also used to test the performance of origami mask specimens. The configuration of the fit test simulator shares many of the features previously described for the filter material tests. The mannequin head located inside the chamber was Cequipped with conductive tubing running from two holes cut into the nostrils to the selection valve. A manual valve and flow meter (model 42260101, TSI, Inc.) were used in place of the flow controller for regulating the sample flow rate of 60 Lpm into the mask. The range of each mask’s expected performance was assessed through: (i) a nominal fit test, or unsealed condition, in which the sample mask was fitted to the mannequin head in an attempt to achieve the best fit; and (ii) the ideal fit test, or sealed condition, where the test mask was sealed to the mannequin head with adhesive tape. The nominal fit test was performed by placing the mask on the mannequin and creating the tightest fit possible but without additional sealing. Since the mannequin head is made from a hard plastic, a mask fit to the head is more likely to leak compared to a similar test performed on normal skin. Thus masks exhibit their lowest filtration efficiencies under these conditions, with leakage also dependent on the flow resistance of the mask material. Once the filtration efficiency and pressure drop were measured, a 2.5 cm wide strip of duct tape was applied to the perimeter of the mask and mannequin head, which effectively sealed the mask to the mannequin face and created conditions for the ideal fit test. The filtration efficiency and pressure drop were then measured to obtain the values under these ideal conditions, both of which were always greater than those obtained during the nominal test.

## Quantitative Fit Test Experiments

Quantitative fit tests were also performed using the ambient aerosol condensation nuclei counter quantitative fit testing protocol approved by the U.S. Occupational Health and Safety Administration (OSHA) and described in Appendix A of Specification 1910.134. These tests allow us to compare our mannequin-based fit test protocol to those commonly applied to industrial and medical masks. Tests were performed using a Portacount Pro+ Model 8038 respirator fit tester (TSI, Inc.). The Portacount reports data as “fit factor,” which is defined as the ambient aerosol concentration divided by the concentration of aerosol inside the mask. Fit factor is the inverse of the transmission efficiency, or 1*/T*_*eff*_, and thus can be directly compared to our mannequin-based filtration efficiency measurements for sealed masks. We confirmed this by comparing the Portacount-derived filtration efficiency to the integrated ideal filtration efficiency obtained by the SMPS using the mannequin-based test protocol described above. Both derived values agreed within measurement uncertainties. Once we established that the Portacount can replicate the integrated filtration efficiencies obtained by our mannequin-based protocol, we performed an OSHA 1910.134 fit test of each mask using a human test subject. These tests were performed using the same NaCl aerosol generation method as that used in our mannequin-based tests and took place in a continuously ventilated 18 m^2^ room. Respiratory fit tests using human subjects differ from our mannequin-based measurements primarily because the former samples both inhaled and exhaled aerosol whereas the latter samples only inhaled aerosol. Thus, depending on the deposition of inhaled particles in the human respiratory system, which has been shown to be significant for the hygroscopic NaCl particles used in this study, ^44^ filtration efficiency calculated from measured fit factor may overestimate face mask filtration efficiency.

## Results and Discussion

We measured the filtration efficiencies of 18 different candidate materials, including multilayered “hybrid” configurations. Plots showing the size-resolved filtration efficiency and quality factor for each sample are presented in the Supporting Information, Figs. S1 - S4. A large range of filtration efficiencies was observed, including nearly 100% for 2-plies of a furnace filter, to as little as 5% for NWPP, which is commonly used in reusable shopping bags. These results show that, in general, materials with higher filtration efficiencies have correspondingly higher pressure drops, which in practice results in a trade-off between efficiency and leakage. Exceptions included materials that are specially designed for filtration applications, including furnace filters and Filti brand filter media.

We selected 15 different single-ply, double-ply, and hybrid configurations of mask materials and conducted a series of fit tests using the mannequin-based protocol presented above and in Fig. 4. For reference, Table 1 in the Methods section provides the name and corresponding label of each mask. Fig. 5 shows representative results from measurements performed on a surgical and KN95 mask with ear loops (for reference) and five masks (masks 1F, iH, H1, H2, H3) that comply with the AFNOR SPEC S76-001 standards with regards to filtration efficiency and pressure drop. The upper solid line of each plot corresponds to the ideal fit test results and the lower solid line corresponds to the nominal test results. At each diameter, the difference between upper and lower lines, represented as a shaded area in Fig. 5, shows the potential effects of leakage from an ill-fitting mask. As discussed above, mask leakage plays a critical role in determining mask efficacy. However, there have been few attempts to quantify this impact. In one study, wherein only filter media were tested and not actual masks, small holes were drilled in the sample tube downstream of the filter in order to simulate leakage by allowing background particles to enter the filtered aerosol flow.^30^ Our approach provides an improved understanding of the impacts of pressure drop on leakage and, in addition, is a better indicator of the actual performance of the mask design. In general, we observed that masks with higher upper bound filtration efficiencies often exhibited larger total efficiency ranges, implying that high efficiency media typically possess high pressure drops, which ultimately results in mask leakage. This phenomenon is clearly evident in the surgical and KN95 reference masks, both of which performed extremely well during the sealed (upper bound) condition (95% and 93% filtration efficiency at 300 nm, respectively), while under-performing during unsealed (lower bound) conditions, (30% for both at 300 nm). For the KN95 mask, we attribute this leakage to the use of ear loops rather than straps that wrap around the head. In our nominal fit tests and in our tests with a human subject, the ear loops were not able to press the mask to the face well enough to eliminate leakage.

**Figure 5:**
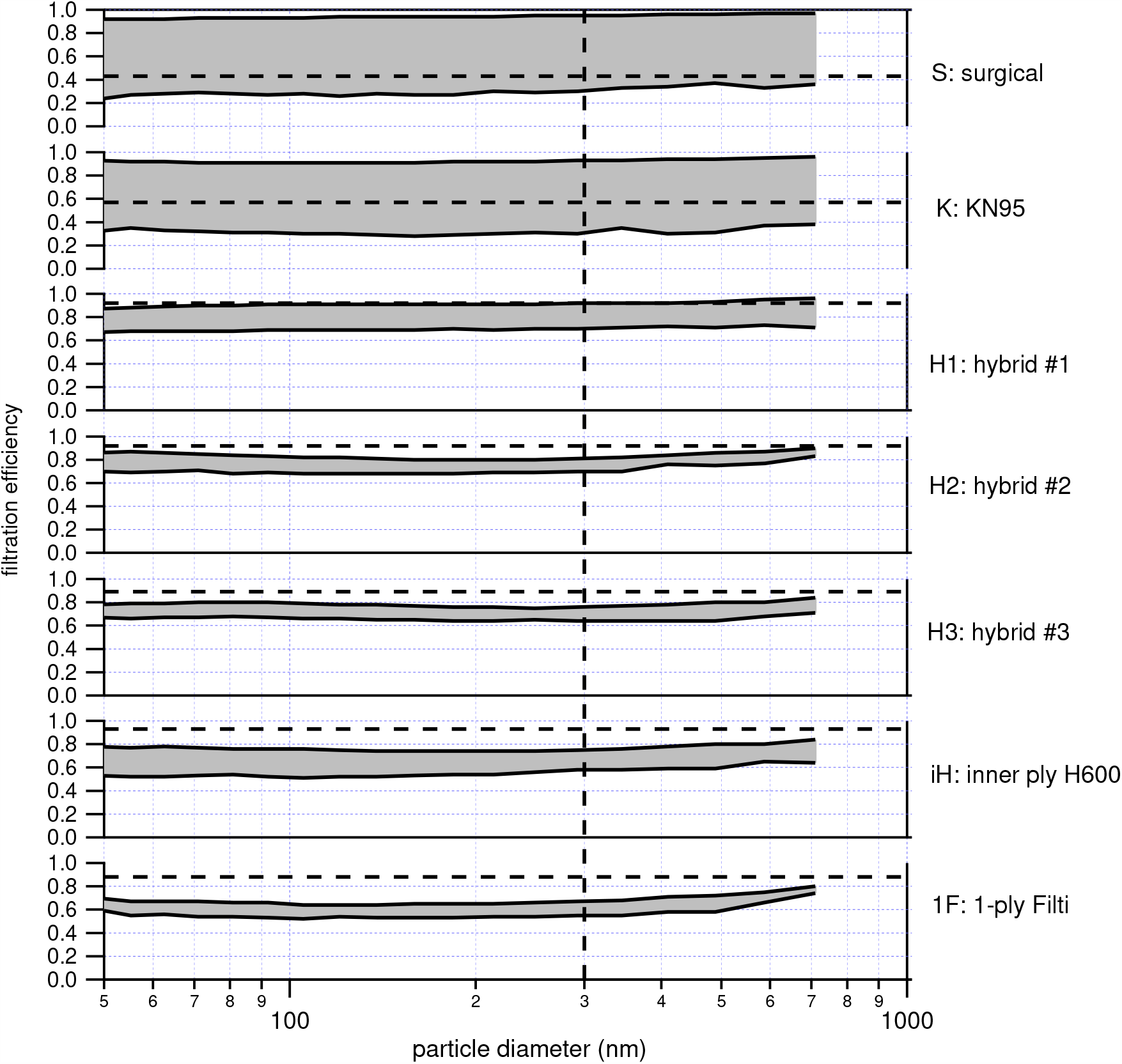
Expected filtration efficiency ranges for origami masks made from different materials at an intake flow rate of 60 Lpm. For each mask, the upper bound filtration efficiency corresponds to the sealed condition, while the lower bound corresponds to the unsealed condition. The dashed vertical line identifies the 300 nm particle diameter and the horizontal dashed line indicates the filtration results obtained from the Portacount respirator fit tester.

**Figure 6:**
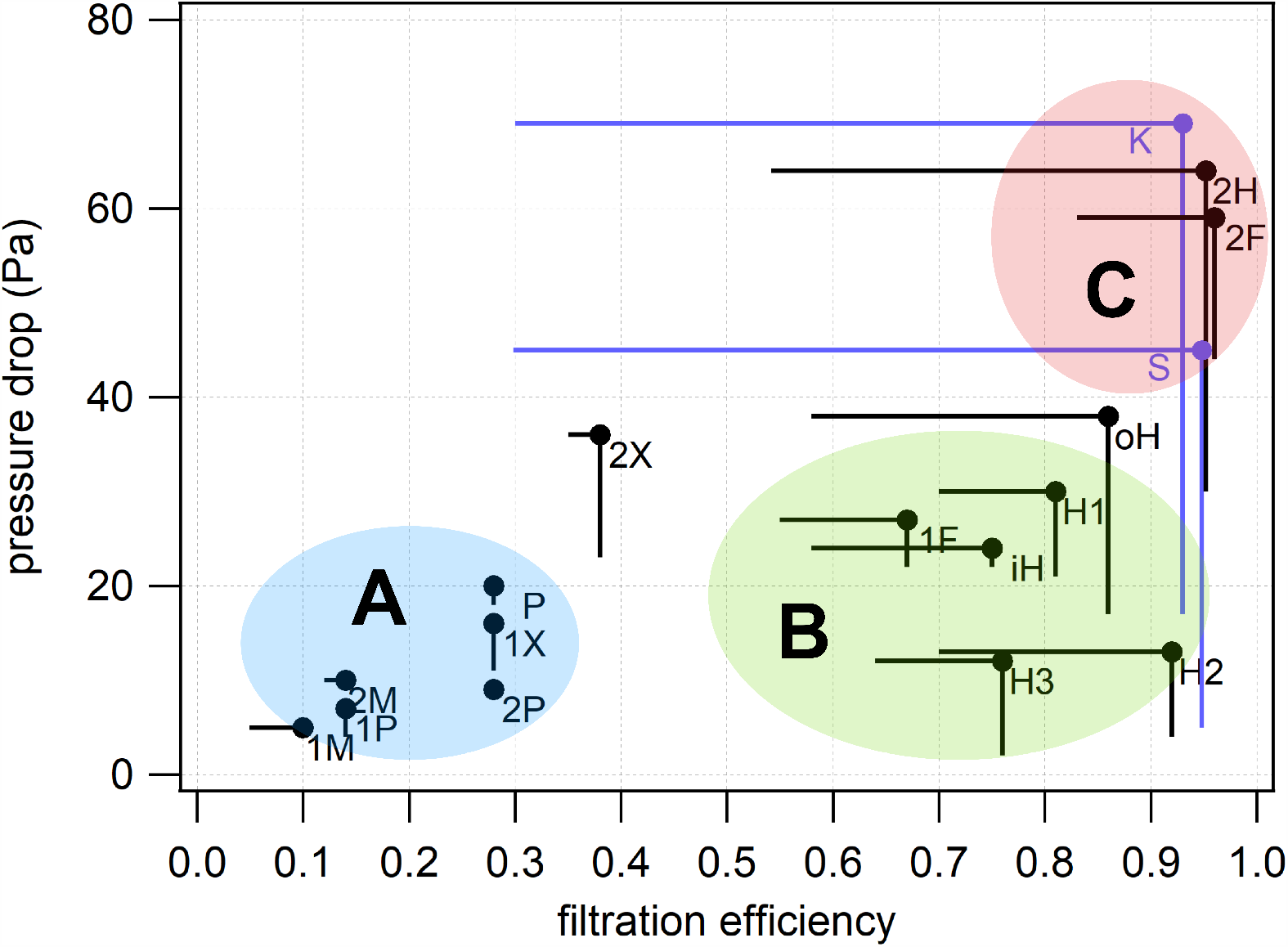
Pressure drop versus filtration efficiency at 300 nm particle diameter for individual origami mask prototypes at an intake flow rate of 60 Lpm. Table 1 in the Methods section lists the material and correpsonding label for each mask. Horizontal lines represent the range of filtration efficiency between the sealed and unsealed test conditions, and vertical lines represent the range of pressure drop between the sealed and unsealed test conditions. Qualitatively, the masks can be clustered into three categories: (**A**) Low efficiency but very low leakage and minimal variability in efficiency. Low leakage means proper mask fit is assured; (**B**) Good filtration and low pressure drop with moderate range of efficiency. (**C**) Excellent filtration but a high pressure drop, meaning a proper face seal might be challenging or impossible for a given mask design.

Similar plots for the other masks that we tested are provided in the Supporting Information (Figs. S5 and S6). These data are grouped according the their filtration performance and pressure drop. The masks plotted in Fig. S5 (masks 1M, 2M, P, 2P, 1X, 2X) show generally lower filtration efficiencies during the sealed condition and relatively small variability when unsealed, suggesting that leakage was minimal. Not surprisingly, these low-efficiency masks had some of the lowest pressure drops measured. In contrast, the masks plotted in Fig. S6 (masks 2H, oH, 2F) show high filtration efficiencies and high variability, suggesting that these masks may experience high leakage in actual use. These masks had some of the highest pressure drops. One exception is the mask made from dual-ply Filti filter (mask 2F), which in spite of having a high filtration efficiency and high pressure drop has relatively low variability, implying lower leakage. In general, our results reveal that filtration performance is dominated by mask fit and that reducing the pressure drop of the filter material attenuates leakage.

We also performed fit tests on a human subject using the quantitative fit test OSHA Specification 1910.134 to provide a point of reference for our measurements of mask performance. Fit factors determined from this protocol were converted to filtration efficiency and plotted for each mask as horizontal dashed lines in Fig. 5 as well as in Figs. S5 and S6. While we do not have a quantitative measurement of NaCl aerosol deposition in the respiratory tract of our human test subject, the overestimate of the filtration efficiency of the fit factor-derived filtration efficiency is clearly seen in Fig. 5 for masks 1M, 2M, P, 2P, 1X, 2X, and also appears to be true for all other origami masks plotted in Figs. S5 and S6. These results suggest that, in actual use, the filtration efficiency of our origami mask is likely close to that measured under ideal conditions in which the mask is sealed to the mannequin. Significantly, the KN95 with ear loops and the surgical mask were the only masks for which the OSHA fit test-derived efficiencies were significantly lower than the mannequin-based ideal efficiency. These masks exhibited significant leakage even with the human test subject, which emphasizes the challenges in minimizing leakage for these mask types.

To better visualize the relationships between filtration efficiency and pressure drop and the variability of each, we plot the range of each mask’s pressure drop (horizontal traces) and filtration efficiency for 300 nm diameter particles (vertical traces) in Fig. 6. This plot clearly shows the different groups represented by Fig. 5 and Figs. S5 and S6. As stated previously, Group (**A**) has low filtration efficiency and low pressure drop and thus horizontal and vertical traces are among the shortest of those in the plot. Group (**C**) has excellent filtration but a high pressure drop making a proper face seal challenging. Not surprisingly, both the surgical and KN95 reference masks cluster in Group (**C**). Group (**B**) has good filtration, low pressure drop and moderate leakage and variability of efficiency. These masks appear to optimize filtration with pressure drop and leakage and are examples of materials that perform well for improvised respiratory protection using this design.

Overall, our results indicate that the origami fabrication technique can achieve a large range of masks with different filtration and pressure drop characteristics. By proper selection of filter media, these masks and can match the filtration performance of commercial respiratory protection while offering benefits of lower pressure drop and minimal leakage.

## Conclusion

We have proposed an origami fabrication technique as a solution for constructing improvised respiratory protection on-demand. We evaluated the filtration properties of variety of readily available candidate materials. We then constructed 15 different origami masks using single-ply, double-ply, and hybrid configurations of a subset of these materials and conducted a series of fit tests in order to characterize the interrelationship between pressure drop, filtration efficiency, and mask leakage. We observed that masks constructed with higher filtration efficiency materials often exhibited a large range of expected efficiencies, implying that airflow through high efficiency media leads to high pressure drops, which increases the likelihood of masks leakage. Our results provide an improved understanding of the impacts of pressure drop on leakage and, in addition, provide a more reliable indicator of the expected performance of a mask design. Importantly, alternative materials such as other fabrics with known filtration efficacy (e.g., see ref. ^29–34^) can be used with the proposed fabrication technique. The origami fabrication procedure represents a flexible solution that can accommodate different facial sizes, and, depending on material choice, realize the full range of filtration properties allowing a user to tune improvised masks to their particular needs. In summary, we have designed a mask that can be produced with minimal cost and labor that optimizes filtration efficiency and ease of breathing, and because this mask can provide greater comfort, compared to some commercial alternatives, it is likely to promote greater mask wearing tolerance and acceptance.

## Data Availability

The data from this study is available upon request.

## Acknowledgements

This research was supported by the Joint Research Fund and Office of Research at the University of California, Irvine through the COVID-19 Basic, Translational and Clinical Research Fund.

## Notes

### Competing Interest Statement

The authors have declared no competing interest.

### Author Declarations

IRB was unnecessary for this study.

